# Ideal Cardiovascular Health in Urban Jamaica: Prevalence Estimates and Relationship to Community Property Value, Household Assets and Educational Attainment

**DOI:** 10.1101/2020.01.12.20017277

**Authors:** Joette A. McKenzie, Novie O. Younger-Coleman, Marshall K. Tulloch-Reid, Ishtar Govia, Nadia R. Bennett, Shelly R. McFarlane, Renee Walters, Damian K. Francis, Karen Webster-Kerr, Andriene Grant, Tamu Davidson, Rainford J. Wilks, David R. Williams, Trevor S. Ferguson

## Abstract

**BACKGROUND:** Ideal cardiovascular health (ICH) is associated with greater longevity and reduced morbidity, but no research on ICH has been conducted in Jamaica. We estimated the prevalence of ICH in urban Jamaica and evaluated associations between ICH and community, household and individual socioeconomic status (SES).

**METHODS:** Cross-sectional study using data from 360 men and 665 women, age ≥20 years in urban Jamaica. ICH was defined as having seven characteristics: current non-smoking, healthy diet, moderate physical activity, and normal body mass index, blood pressure, glucose, and cholesterol. Logistic regression, weighted for survey design, quantified association between the outcome (≥5 ICH characteristics [ICH-5]), and exposure variables (tertiles of community median land value [MLV], tertiles of household assets and educational attainment).

**RESULTS:** Prevalence of ICH (7 characteristics) was 0.51%, while prevalence of ICH-5 was 22.9% (male 24.5%, female 21.5%, p=0.447). In sex-specific multivariable models adjusted for age, education, and household assets, men in the lower tertiles of community MLV had lower odds of ICH-5 compared to men in the upper tertile (lowest tertile: OR 0.33, 95%CI 0.12-0.91, p=0.032; middle tertile: OR 0.46 (0.20-1.04) p=0.062). Women from communities in the lower and middle tertiles of MLV also had lower odds of ICH-5, but association was not statistically significant. Educational attainment was inversely associated with ICH-5 among men and positively associated among women. No significant association was seen for household assets.

**CONCLUSION:** Prevalence of ICH is low in urban Jamaica. Living in poorer communities was associated with lower odds of ICH-5 among men. Higher education was associated with higher odds of ICH-5 among women but lowers odds among men.

## INTRODUCTION

The American Heart Association (AHA), in 2010, recommended that in responding to the public health burden of cardiovascular disease (CVD), the focus should be more on preserving cardiovascular health rather than reducing disease (1). In light of this position, the AHA introduced the concept of ideal cardiovascular health (ICH), defined as having seven characteristics, comprising both health behaviours and health factors, namely: current non-smoking, body mass index <25kg/m^2^, at least 150 minutes of moderate physical activity, healthy diet, normal blood pressure, normal glucose and normal cholesterol levels (1). The concept of ICH is believed to be a more far reaching approach to the promotion of cardiovascular health and is based on data which suggest that persons with low levels of CVD risk factors in mid-life have significantly reduced risk of CVD and total mortality, and longer life expectancy (1-3). Since the publication of the AHA definition of ICH, several studies have sought to estimate prevalence of ICH in various populations. Overall prevalence of ICH (i.e. having all 7 components) is low in most populations, with prevalence estimates often <1% (4-10). Given the low prevalence of all seven ICH components, several investigators have used operational definitions for ICH as having 5-7 components, or performed analyses with the number of ICH components as the outcome variable (5-8, 10).

CVD risk factor prevalence is high in poor urban communities in both low- and high-income countries (11-14). Data from developing countries suggest increasing rates of CVDs in poor urban communities and urbanization has been identified as one of the factors driving the non-communicable disease (NCD) epidemic (11, 15, 16). However, research on cardiovascular health in low- and middle-income countries (LMIC) remains limited and where data exist the focus has been primarily on traditional CVD risk factors such as hypertension or diabetes, and their associated complications.

Poverty is strongly associated with poor health, high morbidity, and shorter life expectancy (17-20). The association between poverty and health has been established as far back as in the 19^th^ century, when Charles Booth reported spatial associations between disease and community location and community poverty (21). Today, poverty related health inequality is seen for both infectious and non-communicable diseases (22). Poverty is associated with increased exposure to unhealthy diets, physical inactivity, air pollution, crime, and increased psychosocial stress – all of which contribute to poorer health (19, 20).

Assessment of socioeconomic status (SES) in LMICs can be challenging. There are few data sources providing information on income, and self-reported income is often limited by high rates of non-response to questionnaire items (23). Most studies have used individual level indicators such as education or occupation as proxies for SES. Increasingly, studies now explore neighbourhood characteristics to assess community SES (24). Property value is also emerging as a useful means of assessing SES (25, 26), sometimes using data from tax related agencies and national censuses (27). In Britain, the Council tax valuation band is used to assess SES (26) along with unimproved land value, household value, overcrowding and household tenure (28-31). Though non-traditional, several of these measures have been shown to be comparable to income and other SES indices, leading to increased use of individual or aggregated property value as a proxy measure of SES (32, 33). Property value can be considered as an indicator of wealth; studies have shown the strong association between property value and other markers of SES previously used as measures of wealth (34, 35).

Jamaica is classified as an upper middle-income country but has a large number of poor ‘inner city’ communities with poverty estimated at 17% (36, 37). Previous studies have documented high prevalence of CVD risk factors in Jamaica, with hypertension prevalence of 25%; diabetes, 8%, hypercholesterolaemia, 12%, obesity, 25% and smoking 15% (38). Additionally, there are reported socioeconomic disparities in the burden of CVDs, with higher age adjusted prevalence of diabetes among men with less education and higher prevalence of hypertension among less educated women (39). To date, there are no published data on the prevalence of ICH or its components or on the relationship between ICH and community or household socioeconomic status.

In this paper we report prevalence estimates of ICH and its individual components in urban Jamaica and evaluate the relationship between ICH and community, household and individual SES. Specifically, we evaluated the association between having five or more ICH characteristics (ICH-5) and community median property value, number of household assets and individual education attainment.

## METHODS

### Study design and data sources

We conducted a cross sectional study using data from the Jamaica Health and Lifestyle Survey 2016-2017 (JHLS III), a national health examination survey of Jamaicans 15 years and older. Trained data collectors used an interviewer-administered questionnaire to obtain data on demographic characteristics, medical history and health behaviours, including dietary practices and physical activity. Blood pressure and anthropometric measurements were done in the participant’s home and followed standardized procedures. A capillary blood sample was collected in the fasting state to measure glucose and cholesterol. For this study, we included urban participants 20 years and older. Data related to the seven ICH characteristics were extracted from the main JHLS III datafile along with demographic data including age, sex, educational attainment and household assets. Ethical approval for JHLS III was granted by the University of the West Indies Ethics Committee and the Ministry of Health Ethics Committee. Additionally, we obtained ethical approval for the current project from the University of the West Indies Ethics Committee. Participants provided written informed consent prior to data collection.

Data on property value were obtained from the National Land Agency [NLA] (http://www.nla.gov.jm/content/background), the governmental body responsible for property valuation in Jamaica. Data were extracted by overlaying a shapefile of all the enumeration districts (EDs) surveyed in the JHLS III to a central database of properties in Jamaica. The shapefile was procured from the Statistical Institute of Jamaica (STATIN), the governmental body responsible for defining ED boundaries. STATIN also provided data on the communities to which each ED belonged based on the community classification by the Social Development Commission (SDC), a government agency with responsibility for community organization and development.

The unimproved land value data for each property within the boundaries of the EDs were added to the shapefile by the NLA and data exported to a spreadsheet. Property values were quoted in Jamaican dollars and parcels classified as either residential or commercial. Data for 25,645 parcels of land from 169 EDs spanning 151 communities were obtained and used to compute median property value for each community.

### Measurement of ICH Variables

Blood pressure was measured using an oscillometric device (Omron 5 series blood pressure monitor, Omron Healthcare, Lake Forest, IL). Measurements were taken in the right arm after the participant had been seated for five minutes and followed standardized procedures developed for the International Collaborative Study of Hypertension in Blacks (40). Weight was measured using a portable digital scale (Tanita HD-351 Digital Weight Scale, Tanita Corporation, Tokyo, Japan) and recorded to the nearest 0.1 kg. Height was measured using a portable stadiometer (Seca 213 Mobile stadiometer, Seca GmbH & Co., Hamburg, Germany) and recorded to the nearest 0.1 cm. Fasting glucose and total cholesterol were measured from a capillary blood sample using a point of care device (SD LipidoCare, Suwon, South Korea). Data on smoking status and physical activity and dietary practices were obtained via interviewer-administered questionnaire.

### Variable creation and definitions

Dichotomous variables for each the seven ICH components were created using definitions from the AHA (1). The means of the second and third of three SBP and DBP measurements were used in the analyses. Normal blood pressure was defined as SBP <120 mmHg and DBP <80 mmHg for persons not on medication for hypertension. BMI was computed as the weight in kilograms divided by the square of height in metres and normal BMI defined as BMI < 25.0 kg/m^2^. Normal glucose was defined as fasting glucose <5.6 mmol/l among persons not on medication for diabetes and normal cholesterol as cholesterol <5.2 mmol/l among persons not on medication for hypercholesterolemia. Persons who reported never smoking or having quit for greater than 12 months were classified as current non-smoker.

Adequate physical activity was defined as engaging in at least 150 minutes of moderate physical activity or 75 minutes of vigorous activity each week. The healthy diet definition was modified based on the data that were collected for JHLS III as we did not have data on whole grain intake. Participants were therefore classified as having a healthy diet if they had three or more of the following dietary characteristics: 1) low salt diet, defined as no added salt at the table and rarely or never eats processed foods; 2) low sugar-sweetened beverage (SSB) consumption, defined as SSB intake < twice per week; 3) adequate fruit and vegetable consumption, defined as intake of fruits or vegetables >= three times per day; and 4) adequate fish consumption, defined as eating fish two or more times per week.

From the seven ICH characteristics we created dichotomous variables for having all seven ICH characteristics (ICH-7), five or more ICH characteristics (ICH-5), four ICH health behaviours (adequate physical activity, non-smoker, normal BMI and healthy diet) and four ICH health factors (normal blood pressure, normal glucose, normal cholesterol and current-non-smoker).

Educational attainment was used as a measure of individual level socioeconomic status, based on participants’ responses to a question on the highest level of education reached. Responses were categorized as ‘less than high school’ for persons reporting no education, elementary/primary school education, or junior secondary school education (up to grade 8), ‘high school’ for participants reporting at least some secondary level education (grades 9-13) and ‘more than high school’ for those reporting at least some post-secondary school education, whether through college, university or vocational training institutions.

Household socioeconomic status was assessed based on participants’ responses to whether they had a list of 22 household assets. The full list of items is show in Table S1 of the supplementary file. We created tertiles of the number of household assets for use in the analyses. Tertile 1 included persons with nine or fewer household assets, tertile 2 included those with 10-12 items and tertile 3 persons with 13-22 items.

Property values for each community were ranked and the median value selected as a summative measure for the community. Community median property values were then ranked (stratified by rural/urban category) and categorized into land value tertiles – lower, middle and upper - and used for further analysis.

### Statistical Analysis

Data analyses were performed using Stata 14.2 software (Stata Corp., College Station, Texas). Survey weights were used to account for survey design and the age and sex distribution of the Jamaican population. Descriptive statistics were computed for outcome and exposure variables and for potential confounders. Differences in characteristics across sex categories, land value tertiles, education categories and household possession categories were compared using t-tests, one-way analysis of variance (ANOVA), or the chi-squared test as appropriate. Prevalence of each ICH component, having all seven ICH components and five or more ICH components were estimated within and across sex and SES categories. There was evidence of sex interaction in the association between ICH-5 and education; we therefore presented sex specific estimates in the models for education and in the multivariable models. Multiple logistic regression was used to quantify the sex-specific association between those with five or more ICH characteristics (ICH-5) and exposure variables (land value tertiles, education category, and household possession categories) with adjustment for age.

The JHLS-III dataset included 1167 urban participants. Analyses for this paper were limited to participants ≥20 years old with available data on age, sex, survey weights and at least one ICH characteristic. This resulted in dataset for analysis with 1025 participants. Statistical significance was set at two-sided *p*<0.05.

Multiple imputation was used to account for missing data. Of the 1025 participants, 435 (42%) had no missing values for twenty-two variables selected for use in the analyses, while 590 participants (58%) had one or more missing values. Except for slightly higher proportion of persons with low salt diet, low sugar-sweetened beverage consumption and more than high school education, there were no other significant differences between participants with missing data and those with non-missing data. The number and proportion of missing values for the main variables included in the analyses are shown in Table S2 of the supplementary file. Proportion of missing values was mostly less than 10%, except for BMI (11%) fasting glucose (26%), fasting cholesterol (28%) and smoking history (32%). While complete case analysis (list-wise deletion of missing data) may have been a valid approach to dealing with missing data, this approach is results in loss of power and potential selection bias; we therefore chose to use multiple imputation to deal with missing data in this paper (41, 42). We generated 32 imputed datasets, such that the number of imputations would be at least equal to the highest proportion of incomplete cases, representing an adaptation of a rule of thumb suggested by White and colleagues (42). Imputation models included all the variables for inclusion in multivariable models. Bivariate and multivariable models were created using Stata’s mi estimate command using the thirty-two multiply imputed data sets; estimates were combined by the software using Rubin’s rules (43, 44).

We also performed sensitivity analyses to assess whether the associations found would remain if assessed differently. The specific additional models included: (1) two-level multilevel models with community as the cluster variable, (2) analysis of covariance model (ANCOVA) using the number of ICH components as the outcome variable, (3) complete case analysis (i.e. model with no imputed values). Findings from these sensitivity analysis models are reported in the supplementary file (Tables S5-S7).

## RESULTS

The analysed sample included 1025 participants (360 men, 665 women) with mean age (± standard deviation [SD]) of 47 ± 17.5 years. There was no sex difference in mean age. Summary statistics for the participant characteristics are shown in Table 1. Males were significantly taller and had a lower BMI than their female counterparts (*p*<0.001). Systolic blood pressure was also significantly higher among males compared to females (133.5 mmHg vs. 129.1 mmHg, *p*<0.01). Three times as many men reported current cigarette smoking, with the prevalence of current smokers being 21.2% among men and 6.1% among women (*p*<0.001). A greater proportion of men had less than a high school education while a higher proportion of females had high school or more than high school educational attainment. There were no significant sex differences in the distribution of household assets or community property value categories.

**Table 1:**
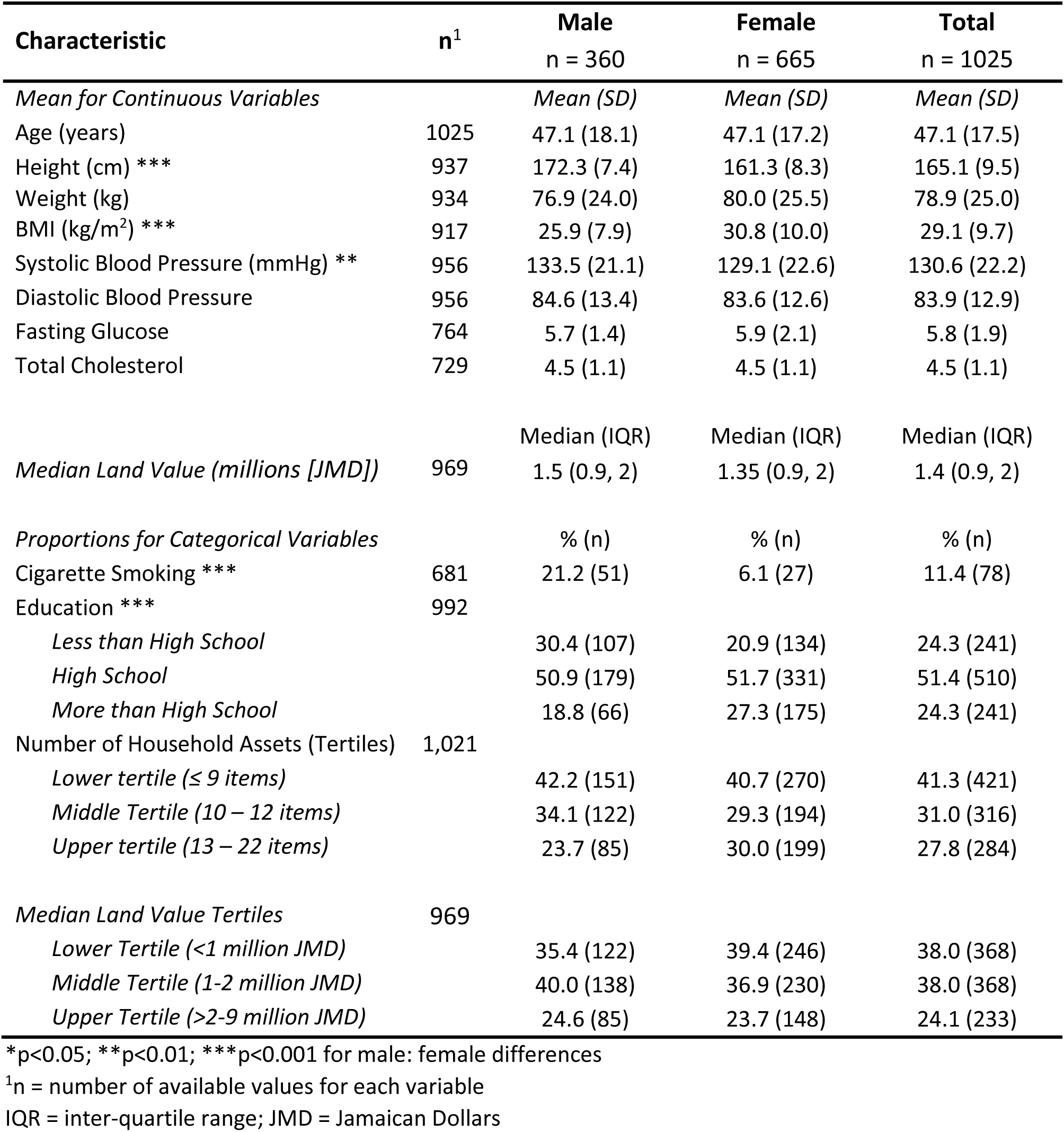
Summary Characteristics of Urban Participants in Jamaica Health and Lifestyle Survey III by sex.

Summary statistics for characteristics stratified by tertiles of community median land value (MLV) are shown in Table S3 of the supplementary file. Except for height, there were no statistically significant differences in mean values for the assessed characteristics by MLV tertile. Height was highest in the middle MLV tertile (*p*<0.05). There was a statistically significant association between MLV tertiles and both education attainment (*p*<0.001) and household assets (*p*<0.001). As expected, persons in the higher MLV tertiles were more likely to have higher education attainment. Similarly, there were higher proportions of persons in the upper tertile of household assets in the higher categories of MLV. Smoking prevalence was highest in the lower MLV tertile, but this did not achieve statistical significance.

The mean number of ICH characteristics and prevalence estimates for overall ICH and other ICH metrics, as well as the individual ICH components for all participants and within sex categories are shown in Table 2. The mean number of ICH characteristics was 3.6 with no significant sex differences. Overall prevalence of ICH (all 7 characteristics, ICH-7) was 0.51%, while the proportion of persons with ≥5 ICH characteristics (ICH-5) was 22.9%. Males had higher prevalence of ICH-5 (24.5%) compared to females (21.5%) but the difference was not statistically significant. We did not compute sex-specific estimates for ICH-7 as there were no men with ICH-7 in the non-imputed data. The proportions of participants having one to seven characteristics is shown in Figure 1. The highest proportion was for four characteristics (29.1%), followed by three characteristics (27.8%). Prevalence of all four health behaviours was 4.1% and was higher in males compared to females, 6.4% vs. 2.0%, *p*=0.014. Prevalence for all four ideal health factors was 14.2% and was higher in females (17.3% vs. 10.6%), but the difference was not statistically significant. Prevalence of individual ICH characteristics ranged from a low of 20% for healthy diet to a high of 87% for non-smoking. For the induvial ICH components, women had significantly higher proportions with normal blood pressure and non-smoking status, while men had higher proportions with normal BMI and adequate physical activity levels. There were no significant sex differences for normal glucose, normal cholesterol and healthy diet.

**Table 2:**
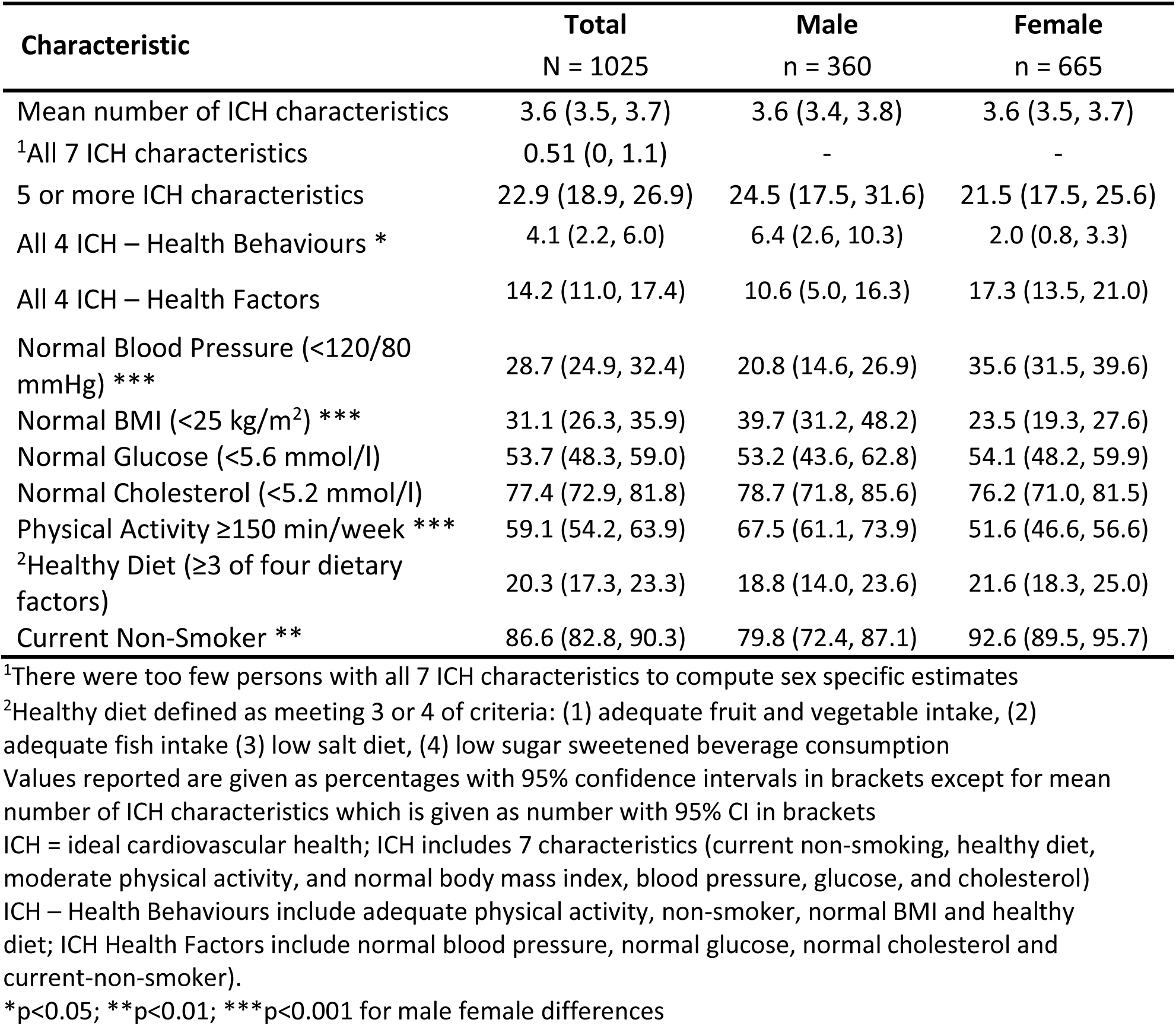
Mean number of Ideal Cardiovascular Health (ICH) Characteristics, Prevalence of overall ICH and Individual ICH Components among of Urban Participants in Jamaica Health and Lifestyle Survey III by sex

| **Characteristic** | **Total**<br>**N = 1025** | **Male**<br>**n = 360** | **Female**<br>**n = 665** |
| --- | --- | --- | --- |
| Mean number of ICH characteristics | 3.6 (3.5, 3.7) | 3.6 (3.4, 3.8) | 3.6 (3.5, 3.7) |
| <sup>1</sup>All 7 ICH characteristics | 0.51 (0, 1.1) | - | - |
| 5 or more ICH characteristics | 22.9 (18.9, 26.9) | 24.5 (17.5, 31.6) | 21.5 (17.5, 25.6) |
| All 4 ICH – Health Behaviours * | 4.1 (2.2, 6.0) | 6.4 (2.6, 10.3) | 2.0 (0.8, 3.3) |
| All 4 ICH – Health Factors | 14.2 (11.0, 17.4) | 10.6 (5.0, 16.3) | 17.3 (13.5, 21.0) |
| Normal Blood Pressure (<120/80 mmHg) *** | 28.7 (24.9, 32.4) | 20.8 (14.6, 26.9) | 35.6 (31.5, 39.6) |
| Normal BMI (<25 kg/m<sup>2</sup>) *** | 31.1 (26.3, 35.9) | 39.7 (31.2, 48.2) | 23.5 (19.3, 27.6) |
| Normal Glucose (<5.6 mmol/l) | 53.7 (48.3, 59.0) | 53.2 (43.6, 62.8) | 54.1 (48.2, 59.9) |
| Normal Cholesterol (<5.2 mmol/l) | 77.4 (72.9, 81.8) | 78.7 (71.8, 85.6) | 76.2 (71.0, 81.5) |
| Physical Activity ≥150 min/week *** | 59.1 (54.2, 63.9) | 67.5 (61.1, 73.9) | 51.6 (46.6, 56.6) |
| <sup>2</sup>Healthy Diet (≥3 of four dietary factors) | 20.3 (17.3, 23.3) | 18.8 (14.0, 23.6) | 21.6 (18.3, 25.0) |
| Current Non-Smoker ** | 86.6 (82.8, 90.3) | 79.8 (72.4, 87.1) | 92.6 (89.5, 95.7) |

<sup>1</sup>There were too few persons with all 7 ICH characteristics to compute sex specific estimates
<sup>2</sup>Healthy diet defined as meeting 3 or 4 of criteria: (1) adequate fruit and vegetable intake, (2) adequate fish intake (3) low salt diet, (4) low sugar sweetened beverage consumption
Values reported are given as percentages with 95% confidence intervals in brackets except for mean number of ICH characteristics which is given as number with 95% CI in brackets
ICH = ideal cardiovascular health; ICH includes 7 characteristics (current non-smoking, healthy diet, moderate physical activity, and normal body mass index, blood pressure, glucose, and cholesterol)
ICH – Health Behaviours include adequate physical activity, non-smoker, normal BMI and healthy diet; ICH Health Factors include normal blood pressure, normal glucose, normal cholesterol and current-non-smoker).
\*p<0.05; \*\*p<0.01; \*\*\*p<0.001 for male female differences

**Figure 1:**
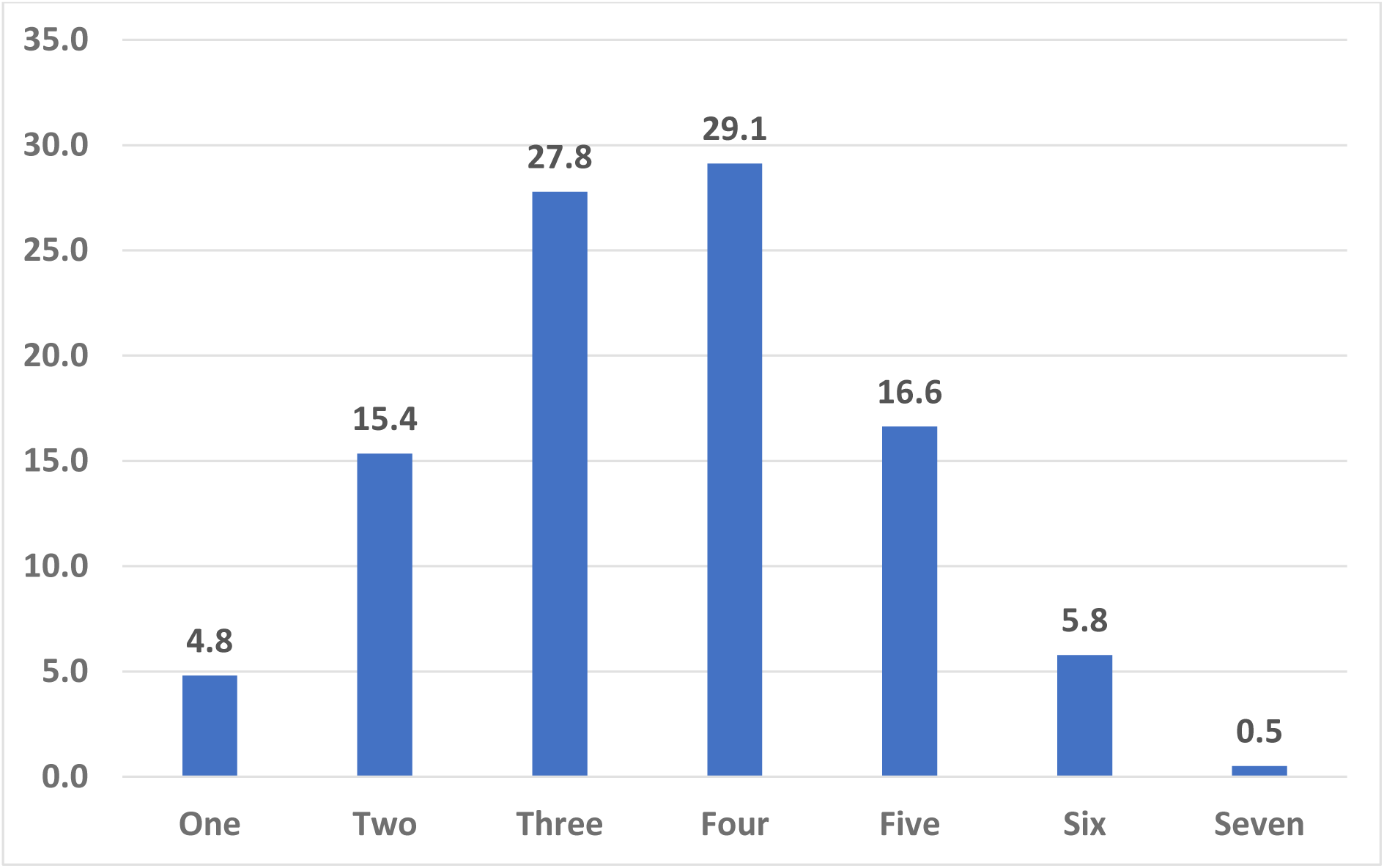
Proportion (%) of Participants with 1-7 ICH components.

Summary statistics for the ICH characteristics described above, stratified by levels of community property value, household assets and individual educational attainment are shown in Tables S4A – S4C in the supplementary file. There were no significant associations between mean number of ICH characteristics across MLV tertiles. Prevalence of ICH-5 was higher in the upper and middle tertiles of MLV but the differences were not statistically significant. Prevalence of all four ideal health behaviours was also higher in the middle and upper tertiles of MLV, this time achieving statistical significance for the lower MLV tertile when compared to the upper MLV tertile. Prevalence of ideal health factors was not significantly different across MLV tertiles. For the individual ICH characteristics, significant differences were seen for normal cholesterol and healthy diet. Prevalence of normal cholesterol was higher among those in the lower tertile of MLV, while prevalence of healthy diet was highest among those in the upper MLV tertile.

There were no significant differences between mean number of ICH characteristics and tertiles of household assets (Table S4B in supplementary file). Prevalence of ICH-5 was highest in the middle tertile of household assets and lowest in the lower tertile, but the differences were not statistically significant. There were no significant differences in the prevalence of ideal health behaviours or ideal health factors. Proportions for non-smokers and those with healthy diet were significantly higher among those in the upper tertile of household assets, while adequate physical activity was highest in the middle tertile. There were no differences in prevalence of normal BMI, normal glucose, normal BP or normal cholesterol across categories of household assets.

For educational attainment we report sex-specific estimates because there was evidence of sex interaction in the relationship between ICH-5 and education categories (Table S4C in supplementary file). Among males, mean number of ICH characteristics and proportion of ICH-5 was highest in the middle education (high school) category, with the difference in proportion for ICH-5 approaching statistical significance (*p*=0.061). Prevalence of ideal health behaviours and ideal health factors was also highest among those with high school education, but again not statistically significant. Among females, there were statistically significant differences in the mean number of ICH characteristics and prevalence of ICH-5, with values highest among women with more than high school education and lowest among those with less than high school education. Similar associations were seen for ideal health factors, while for ideal health behaviours the lowest prevalence was in the high school category. With regard to the individual ICH characteristics, only healthy diet showed significant differences by education category among men, with the prevalence of healthy diet being highest among men with less than high school education. Current non-smoking was highest among men with more than high school education, with the difference approaching statistical significance (*p*=0.063 for more than high school vs. less than high school). Among females, significant associations were found for normal BP, normal glucose, normal cholesterol and healthy diet. Prevalence was highest among women with more than high school education and lowest among those with less than high school education for normal BP and normal glucose. For healthy diet however, prevalence was lowest among those with high school education, while for normal cholesterol, the highest prevalence was among women with high school education.

Bivariate models yielding odds ratios for ICH-5 comparing levels of explanatory variables and potential confounders are shown in Table 3. We report sex-specific models, given the sex-interaction between ICH-5 and education categories. Age was inversely associated with ICH-5 for both males and females. Persons in the lower MLV tertiles were less likely to have ICH-5 but this was not statistically significant in the bivariate models. Women with lower education were less likely to have ICH-5 (p<0.05) while men with less education had non-significant higher odds of ICH-5 in bivariate models. No significant associations were seen for tertiles of household assets.

**Table 3:**
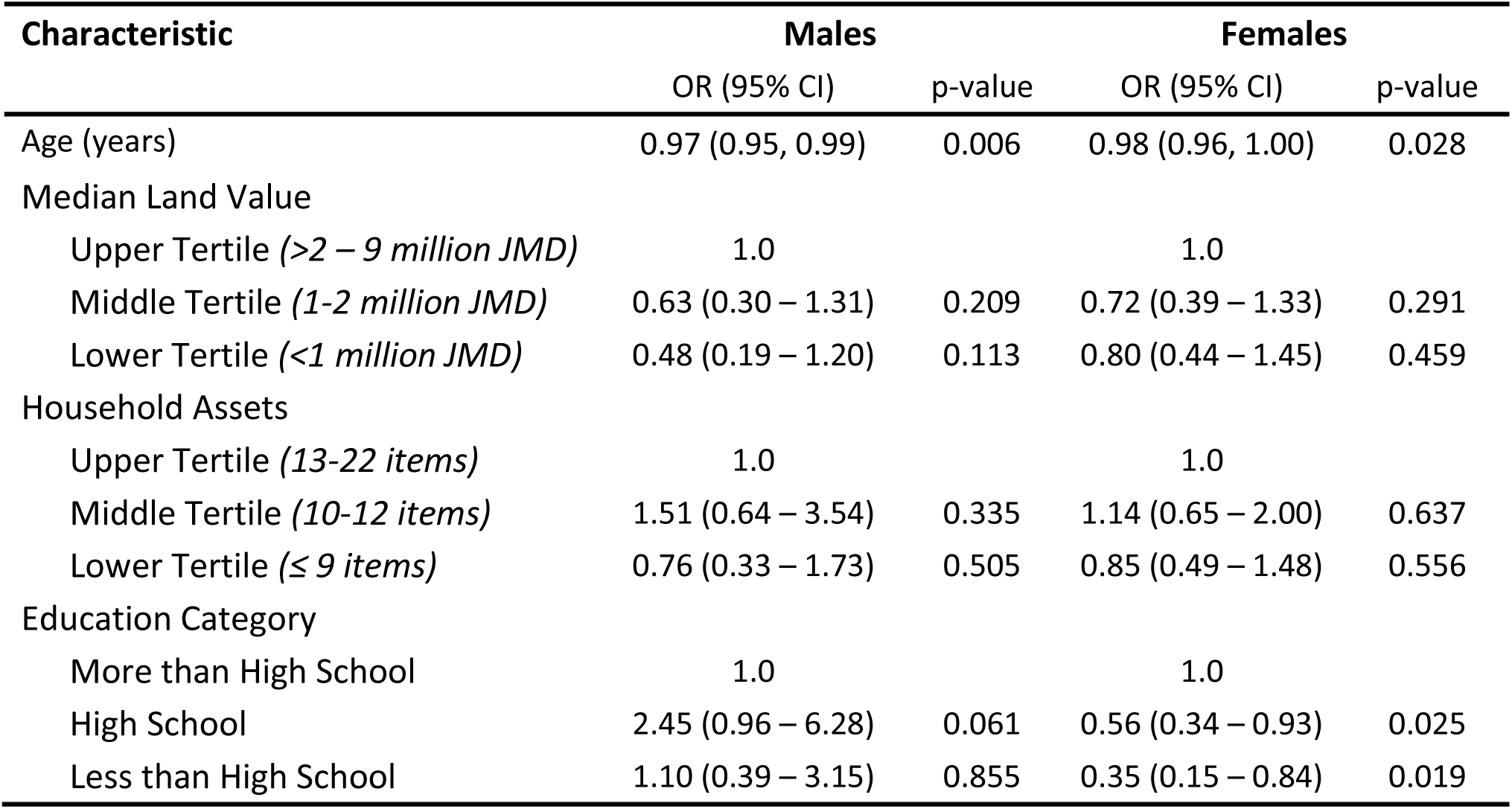
Odds Ratio for ≥5 Ideal Cardiovascular Health Characteristics among of Urban Participants in Jamaica Health and Lifestyle Survey (sex-specific bivariate models)

| Characteristic | Males |  | Females |  |
| --- | --- | --- | --- | --- |
|  | OR (95% CI) | p-value | OR (95% CI) | p-value |
| Age (years) | 0.97 (0.95, 0.99) | 0.006 | 0.98 (0.96, 1.00) | 0.028 |
| Median Land Value |  |  |  |  |
| Upper Tertile (>2 – 9 million JMD) | 1.0 |  | 1.0 |  |
| Middle Tertile (1-2 million JMD) | 0.63 (0.30 – 1.31) | 0.209 | 0.72 (0.39 – 1.33) | 0.291 |
| Lower Tertile (<1 million JMD) | 0.48 (0.19 – 1.20) | 0.113 | 0.80 (0.44 – 1.45) | 0.459 |
| Household Assets |  |  |  |  |
| Upper Tertile (13-22 items) | 1.0 |  | 1.0 |  |
| Middle Tertile (10-12 items) | 1.51 (0.64 – 3.54) | 0.335 | 1.14 (0.65 – 2.00) | 0.637 |
| Lower Tertile (≤ 9 items) | 0.76 (0.33 – 1.73) | 0.505 | 0.85 (0.49 – 1.48) | 0.556 |
| Education Category |  |  |  |  |
| More than High School | 1.0 |  | 1.0 |  |
| High School | 2.45 (0.96 – 6.28) | 0.061 | 0.56 (0.34 – 0.93) | 0.025 |
| Less than High School | 1.10 (0.39 – 3.15) | 0.855 | 0.35 (0.15 – 0.84) | 0.019 |

Table 4 shows the results for our final multivariable model with ICH-5 as the outcome variable. Again, we report sex-specific estimates. Models include the three SES variables and age. We did not include the other cardiovascular risk variables as potential confounders since they were included in the determination of ICH. Men in the lowest tertile of community MLV had 67% lower odds of ICH-5 compared to those in the upper tertile (*p*=0.032), while men in the middle tertile had 54% lower odds of ICH-5 (*p*=0.062). Women from communities in the lower and middle tertiles of MLV were also less likely to have ICH-5; 28% and 30% reduction in the odds of ICH, respectively, but this did not achieve statistical significance. Educational attainment was inversely associated with ICH-5 among men (OR 3.16, *p*=0.079; and OR 3.03, *p*=0.028, for less than high school and high school, respectively, when compared to more than high school education). In contrast, there was a positive association between education level and ICH among women, with lower odds of ICH among those with less education (OR 0.40, *p*=0.070 for less than high school, and OR 0.51, *p*=0.016, for high school, compared to more than high school education). There were no significant associations between the number of household assets and ICH-5 for either men or women.

**Table 4:** Odds Ratio for ≥5 Ideal Cardiovascular Health Characteristics among of Urban Participants in Jamaica Health and Lifestyle Survey (multivariate models)

| Characteristic | Males |  | Females |  |
| --- | --- | --- | --- | --- |
|  | OR (95% CI) | p-value | OR (95% CI) | p-value |
| Median Land Value |  |  |  |  |
| Upper Tertile (>2 – 9 million JMD) | 1.0 |  | 1.0 |  |
| Middle Tertile (1-2 million JMD) | 0.46 (0.20 – 1.04) | 0.062 | 0.70 (0.37 – 1.32) | 0.271 |
| Lower Tertile (<1 million JMD) | 0.33 (0.12 – 0.91) | 0.032 | 0.72 (0.38 – 1.34) | 0.295 |
| Household Assets |  |  |  |  |
| Upper Tertile (13-22 items) | 1.0 |  | 1.0 |  |
| Middle Tertile (10-12 items) | 1.58 (0.68 – 4.09) | 0.340 | 1.41 (0.69 – 2.88) | 0.346 |
| Lower Tertile (≤ 9 items) | 0.81 (0.31 – 2.09) | 0.658 | 1.16 (0.63 – 2.13) | 0.635 |
| Education Category |  |  |  |  |
| More than High School | 1.0 |  | 1.0 |  |
| High School | 3.03 (1.13 – 8.09) | 0.028 | 0.51 (0.29 – 0.88) | 0.016 |
| Less than High School | 3.16 (0.87 – 11.41) | 0.079 | 0.40 (0.15 – 1.07) | 0.070 |
| Age (years) | 0.96 (0.93 – 0.99) | 0.004 | 0.98 (0.95 – 1.00) | 0.092 |

Separate models created for males and female participants in logistic regression models weighted for survey design using multiple imputation to account for missing data. The model for males included 360 participants while the model for females included 665 participants.

For then sensitivity analysis, we re-ran our final model using the following variations: (1) using two-level multilevel models, with community as the cluster variable, (2) using the number of ICH components as the outcome variable (ANCOVA model), (3) using only complete cases (i.e. model with no imputed values). These models are shown in Table S5, S6 and S7 in the supplementary file. The findings produced were generally similar to the those from the final model shown in Table 4. When multi-level logistic regression models were used with ICH-5 as the outcome variable, associations were similar but somewhat attenuated, with some associations no longer statistically significant. In the ANCOVA models, men showed statistically significant inverse associations between number of ICH characteristics and MLV tertile and positive association with education category. Among women, only education attainment showed a significant inverse association in the ANCOVA models. In the complete case-analysis (Table S7) the direction of the associations was again similar to the final model using multiple imputations (see Table 4) but were less precise with wider confidence intervals and sometimes not achieving statistical significance.

## DISCUSSION

In this study we have found that the prevalence of ICH is low in urban Jamaica, with only 0.5% of the population ≥20 years having all seven ICH characteristics and 23% having ≥5 characteristics. Prevalent ICH-5 was associated with community SES measured by median property value. Poorer community SES (lower median land value) was associated with lower odds of ICH-5 among both men and women, but the association was statistically significant among men only. Men with less education had higher odds of ICH-5 while women with less education were less likely to have ICH-5. There were no significant association with number of household assets for either men or women.

The low prevalence of ICH as reported in this study has been seen in several other studies, with prevalence of all seven ICH components being <1% in most studies and prevalence of five or more characteristics ranging between 4% and 37% (6-9, 45-56). A summary of estimates from published studies is shown in Table S8 in the supplementary file. The highest prevalence of 5 or more ICH characteristics was 37% among French-speaking adults in Quebec (46), while the lowest prevalence was among Black participants in the Jackson Heart Study (45). Prevalence varied with age and sex and was generally lower among men and among older persons. There were also some racial differences in the prevalence of ICH, with Blacks having lower prevalence when compared to Whites (49, 56, 57).

With regard to SES, several studies also reported socioeconomic differences in the prevalence of ICH. The majority of studies showed a direct relationship where higher SES was associated with higher prevalence of ICH or mean number of ICH characteristics (46, 47, 50, 51, 53, 57, 58). An analysis of data from the Multi-Ethnic Study of Atherosclerosis (MESA) found that higher income and education were associated with higher prevalence of ICH and that higher neighbourhood physical, social and economic status were also associated higher prevalence of ICH (57). In contrast to the findings in other studies, a study from Peru found that persons in the middle and highest tertile of a wealth index (derived from an aggregation of assets and household facilities were less likely to have ≥5 ICH components (50). We did not find any studies published that specifically assessed the association between property value and ICH.

In this study we used the median land value of the communities as a measure of community SES and found that community SES was inversely associated with prevalence of ≥5 ICH components. Although we did not find similar studies in the literature, our findings were in keeping with those of other studies which found people who live in lower SES urban neighbourhoods were more likely to have a greater burden of cardiovascular disease (59, 60). The findings also corroborate the association between council tax valuation bands (a measure of property value in the United Kingdom) and cardiovascular disease risk factors such as smoking, poor diet and obesity (26). Additionally, our study is consistent with another study which showed that relative location factor (derived from residential property value) was associated with five cardiometabolic risk factors (central obesity, hypertriglyceridemia, reduced HDL, diabetes/diabetic risk, hypertension) and overall cardiometabolic risk score in Australia (34). Our study demonstrates that using property value data from government agencies such as the NLA can circumvent the challenges encountered when collecting epidemiological data used to estimate SES.

While education attainment level and ICH-5 had a positive relationship for women, the reverse was true for men. Looking at the sex-specific proportions for MLV, it was noted that more men were found to live in middle and upper MLV communities than women. It was also found that women were more likely to have higher education than their male counterparts. It is possible that the lower prevalence of ICH-5 among men with higher education may be driven by a higher prevalence of obesity and the metabolic syndrome in men of higher SES which has been previously reported by our team (39, 61). As shown in Table S4C in the supplementary file, men with higher education had lower prevalence of normal BMI, physical activity and healthy diet. These may reflect more sedentary work activities and less time for physical activity and preparing healthy meals. The opposite may often true for higher SES women, who are more likely to be health conscious and thus pay greater attention to diet and participate in leisure time physical activity such as going to the gym.

In this study we did not find an association between household assets and ICH. In a previous study we found a significant inverse association between elevated blood pressure and hypertension for women only (62). Similar to our findings, the Prospective Urban Rural Epidemiologic (PURE) study found that education was more strongly associated with cardiovascular disease than was household wealth based on an index which included ownership of assets and housing characteristics (63).

There were some limitations to this study. First, a complete picture of property value includes that of the house and associated lands, rather than the unimproved land value which was used in this study. It is however expected that land values will be highly correlated with house and total property value and when used for classification into categories as used in this study will correctly classify community SES. Additionally, we did not consider housing tenure, or property ownership, but we believe that the contextual effects of community SES will still be relevant whether or not one owns the property in which he or she resides. The study was also limited by missing data for a number of variables. We were, however, able to use multiple imputation to fill in missing data and perform analyses using the multiple imputed data. This approach reduces potential bias when compared to a complete case analysis and improves the power of the study to show significant associations (42, 64). We also performed sensitivity analyses using only complete cases and found that the direction of association and effect sizes were generally similar, thus indicating that the use of multiple imputation, while improving the power of the study, did not alter the inferences made in the study. Finally, the cross-sectional design limits our ability to make causal inference; however, the findings make a useful contribution to our understanding of the associations between SES and cardiovascular health.

Strengths of the study include the fact that it is was a population-based and that it was designed to be representative of the general urban population in Jamaica. Estimates were weighted to account for the complex survey design, thus ensuring that we can generalize the findings to urban Jamaica. Additionally, the study used standardized methods for measurements and data collection in order to ensure high data quality.

## CONCLUSION

Among urban Jamaicans, prevalence of all seven ICH characteristics was less than 1%, while only 23% of the population had five or more characteristics. Persons who live in poorer communities were less likely to attain ICH-5. Higher education was associated with higher odds of ICH-5 among women but lowers odds among men. We recommend that SES be considered when planning initiatives to promote cardiovascular health especially among urban-dwelling populations. Our data suggest that these efforts may be most effective among Jamaican men and women from poorer communities and among men with higher education.

## Data Availability

Data are available upon request to the corresponding author.

## Acknowledgements

This research was funded by the Bernard Lown Scholars in Cardiovascular Health Program of the Department of Global Health and Population of the Harvard T.H. Chan School of Public Health. The JHLS-III was funded by the Ministry of Health (Jamaica) and the National Health Fund (Jamaica)

We would like to thank the Harvard T.H. Chan School of Public Health Lown Scholars Program for the grant support. We also thank the National Land Agency (NLA) for providing the property value data and the Statistical Institute of Jamaica (STATIN) for providing the shapefile. We also would like to thank the Ministry of Health and Wellness and the National Health Fund Jamaica, which provided funding for the Jamaica Health and Lifestyle Survey III in 2016/17. We would also like to thank the geoinformation unit from both STATIN and NLA. Thanks to the JHLS-III field staff and the CAIHR/ERU administrative staff and the unit driver for their support.

## Notes

### Competing Interest Statement

The authors have declared no competing interest.

## REFERENCES

1. Lloyd-Jones DM, Hong Y, Labarthe D, Mozaffarian D, Appel LJ, Horn LV, et al. Defining and Setting National Goals for Cardiovascular Health Promotion and Disease Reduction. Circulation. 2010;121(4):586–613.

2. Lloyd-Jones DM, Leip EP, Larson MG, D’Agostino RB, Beiser A, Wilson PW, et al. Prediction of lifetime risk for cardiovascular disease by risk factor burden at 50 years of age. Circulation. 2006;113(6):791–8.

3. Stamler J, Stamler R, Neaton JD, Wentworth D, Daviglus ML, Garside D, et al. Low risk-factor profile and long-term cardiovascular and noncardiovascular mortality and life expectancy: findings for 5 large cohorts of young adult and middle-aged men and women. Jama. 1999;282(21):2012–8.

4. Bi Y, Jiang Y, He J, Xu Y, Wang L, Xu M, et al. Status of cardiovascular health in Chinese adults. J Am Coll Cardiol. 2015;65(10):1013–25.

5. Folsom AR, Yatsuya H, Nettleton JA, Lutsey PL, Cushman M, Rosamond WD. Community prevalence of ideal cardiovascular health, by the American Heart Association definition, and relationship with cardiovascular disease incidence. J Am Coll Cardiol. 2011;57(16):1690–6.

6. Lu Y, Shen S, Qi H, Fang N, Li F, Wang L, et al. Prevalence of ideal cardiovascular health in southeast Chinese adults. Int J Cardiol. 2015;184:385–7.

7. Moghaddam MM, Mohebi R, Hosseini F, Lotfaliany M, Azizi F, Saadat N, et al. Distribution of ideal cardiovascular health in a community-based cohort of Middle East population. Annals of Saudi medicine. 2014;34(2):134–42.

8. Zeng Q, Dong SY, Song ZY, Zheng YS, Wu HY, Mao LN. Ideal cardiovascular health in Chinese urban population. Int J Cardiol. 2013;167(5):2311–7.

9. Velasquez-Melendez G, Felisbino-Mendes MS, Matozinhos FP, Claro R, Gomes CS, Malta DC. Ideal cardiovascular health prevalence in the Brazilian population - National Health Survey (2013). Revista brasileira de epidemiologia = Brazilian journal of epidemiology. 2015;18 Suppl 2:97–108.

10. Wu S, Huang Z, Yang X, Zhou Y, Wang A, Chen L, et al. Prevalence of ideal cardiovascular health and its relationship with the 4-year cardiovascular events in a northern Chinese industrial city. Circulation Cardiovascular quality and outcomes. 2012;5(4):487–93.

11. de-Graft Aikins A, Kushitor M, Koram K, Gyamfi S, Ogedegbe G. Chronic non-communicable diseases and the challenge of universal health coverage: insights from community-based cardiovascular disease research in urban poor communities in Accra, Ghana. BMC Public Health. 2014;14 Suppl 2:S3.

12. Nayyar D, Hwang SW. Cardiovascular Health Issues in Inner City Populations. The Canadian journal of cardiology. 2015;31(9):1130–8.

13. De Maio FG. Understanding chronic non-communicable diseases in Latin America: towards an equity-based research agenda. Globalization and health. 2011;7:36.

14. Prabhakaran D, Jeemon P, Reddy KS. Commentary: Poverty and cardiovascular disease in India: do we need more evidence for action? Int J Epidemiol. 2013;42(5):1431–5.

15. Mberu B, Wamukoya M, Oti S, Kyobutungi C. Trends in Causes of Adult Deaths among the Urban Poor: Evidence from Nairobi Urban Health and Demographic Surveillance System, 2003-2012. Journal of urban health : bulletin of the New York Academy of Medicine. 2015;92(3):422–45.

16. World Health Organization. Noncommunicable Diseases Fact Sheet 2015 [Available from: http://www.who.int/mediacentre/factsheets/fs355/en/.

17. Chetty R, Stepner M, Abraham S, Lin S, Scuderi B, Turner N, et al. The Association Between Income and Life Expectancy in the United States, 2001-2014Association Between Income and Life Expectancy in the United StatesAssociation Between Income and Life Expectancy in the United States. JAMA. 2016;315(16):1750–66.

18. Glymour M, Avendano M, Kawachi I. Socioeconomic Status and Health. In: Berkman L, Kawachi I, Glymour M, editors. Social Epidemiology. 2nd edition ed. New York: Oxford University Press; 2014. p. 17–62.

19. Price JH, Khubchandani J, Webb FJ. Poverty and Health Disparities: What Can Public Health Professionals Do? Health promotion practice. 2018;19(2):170–4.

20. Shaw M, Dorling D, Davey Smith G. Poverty, social exclusion, and minorities. In: Marmot M, Wilkinson RG, editors. Social Determinants of Health. Second edition ed. Oxford Scholarship Online: Oxford University Press; 2005.

21. Vaughan L. Charles Booth and the mapping of poverty. Mapping Society. The Spatial Dimensions of Social Cartography: UCL Press; 2018. p. 61–92.

22. Wilkinson RG, Marmot M. Social determinants of health: the solid facts: World Health Organization; 2003.

23. Wilks R, Younger N, Mullings J, Zohoori N, Figueroa P, Tulloch-Reid M, et al. Factors affecting study efficiency and item non-response in health surveys in developing countries: the Jamaica national healthy lifestyle survey. BMC medical research methodology. 2007;7(1):13.

24. Diez Roux AV, Mair C. Neighborhoods and health. Annals of the New York Academy of Sciences. 2010;1186(1):125–45.

25. Connolly S, O’reilly D, Rosato M. House value as an indicator of cumulative wealth is strongly related to morbidity and mortality risk in older people: a census-based cross-sectional and longitudinal study. International Journal of Epidemiology. 2009;39(2):383–91.

26. Fone DL, Dunstan F, Christie S, Jones A, West J, Webber M, et al. Council tax valuation bands, socio-economic status and health outcome: a cross-sectional analysis from the Caerphilly Health and Social Needs Study. BMC public health. 2006;6:115-.

27. Rehm CD, Moudon AV, Hurvitz PM, Drewnowski A. Residential property values are associated with obesity among women in King County, WA, USA. Social science & medicine (1982). 2012;75(3):491–5.

28. Galobardes B, Shaw M, Lawlor DA, Lynch JW, Davey Smith G. Indicators of socioeconomic position (part 1). Journal of epidemiology and community health. 2006;60(1):7–12.

29. Lewis G, Bebbington P, Brugha T, Farrell M, Gill B, Jenkins R, et al. Socioeconomic status, standard of living, and neurotic disorder. The Lancet. 1998;352(9128):605–9.

30. Jackson E, Kupke V, Rossini P. The relationship between socio-economic indicators and residential property values in Darwin: Pacific Rim Real Estate Society; 2007.

31. Corvalán C, Amigo H, Bustos P, Rona RJ. Socioeconomic risk factors for asthma in Chilean young adults. American journal of public health. 2005;95(8):1375–81.

32. Ware JK. Property Value as a Proxy of Socioeconomic Status in Education. Education and Urban Society. 2019;51(1):99–119.

33. Leonard T, Powell-Wiley TM, Ayers C, Murdoch JC, Yin W, Pruitt SL. Property Values as a Measure of Neighborhoods: An Application of Hedonic Price Theory. Epidemiology (Cambridge, Mass). 2016;27(4):518–24.

34. Coffee NT, Lockwood T, Hugo G, Paquet C, Howard NJ, Daniel M. Relative residential property value as a socio-economic status indicator for health research. International Journal of Health Geographics. 2013;12(1):22.

35. Moudon AV, Cook AJ, Ulmer J, Hurvitz PM, Drewnowski A. A neighborhood wealth metric for use in health studies. American journal of preventive medicine. 2011;41(1):88–97.

36. Henry-Lee A. The nature of poverty in the garrison constituencies in Jamaica. Environment and Urbanization. 2005;17(2):83–99.

37. The World Bank. The World Bank in Jamaica: The World Bank; 2019 [Available from: https://www.worldbank.org/en/country/jamaica/overview#1.

38. Ferguson TS, Francis DK, Tulloch-Reid MK, Younger NO, McFarlane SR, Wilks RJ. An update on the burden of cardiovascular disease risk factors in Jamaica: findings from the Jamaica Health and Lifestyle Survey 2007-2008. The West Indian medical journal. 2011;60(4):422–8.

39. Ferguson T, Younger-Coleman N, Tulloch-Reid M, Hambleton I, MacLeish M, Hennis A, et al. Educational Health Disparities in Cardiovascular Disease Risk Factors in Jamaica: Findings from the Jamaica Health and Lifestyle Surveys. The West Indian medical journal. 2013;62(Suppl 2):31.

40. Ataman SL, Cooper R, Rotimi C, McGee D, Osotimehin B, Kadiri S, et al. Standardization of blood pressure measurement in an international comparative study. Journal of clinical epidemiology. 1996;49(8):869–77.

41. Nguyen CD, Carlin JB, Lee KJ. Model checking in multiple imputation: an overview and case study. Emerging Themes in Epidemiology. 2017;14(1):8.

42. White IR, Royston P, Wood AM. Multiple imputation using chained equations: Issues and guidance for practice. Statistics in Medicine. 2011;30(4):377–99.

43. Marshall A, Altman DG, Holder RL, Royston P. Combining estimates of interest in prognostic modelling studies after multiple imputation: current practice and guidelines. BMC Med Res Methodol. 2009;9:57.

44. StataCorp. Stata Multiple Imputation Reference Manual Release 14. College Station, TX: StataCorp LLC.; 2015.

45. Djousse L, Petrone AB, Blackshear C, Griswold M, Harman JL, Clark CR, et al. Prevalence and changes over time of ideal cardiovascular health metrics among African-Americans: the Jackson Heart Study. Preventive medicine. 2015;74:111–6.

46. Harrison S, Couillard C, Robitaille J, Vohl MC, Belanger M, Desroches S, et al. Assessment of the American Heart Association’s “Life’s simple 7” score in French-speaking adults from Quebec. Nutrition, metabolism, and cardiovascular diseases : NMCD. 2019;29(7):684–91.

47. Jankovic J, Davidovic M, Bjegovic-Mikanovic V, Jankovic S. Status of cardiovascular health in the Republic of Serbia: Results from the National Health Survey. PLoS One. 2019;14(3):e0214505.

48. Kim JI, Sillah A, Boucher JL, Sidebottom AC, Knickelbine T. Prevalence of the American Heart Association’s “ideal cardiovascular health” metrics in a rural, cross-sectional, community-based study: the Heart of New Ulm Project. Journal of the American Heart Association. 2013;2(3):e000058.

49. Short VL, Gamble A, Mendy V. Racial differences in ideal cardiovascular health metrics among Mississippi adults, 2009 Mississippi Behavioral Risk Factor Surveillance System. Preventing chronic disease. 2013;10:E194.

50. Benziger CP, Zavala-Loayza JA, Bernabe-Ortiz A, Gilman RH, Checkley W, Smeeth L, et al. Low prevalence of ideal cardiovascular health in Peru. Heart. 2018;104(15):1251–6.

51. Empana JP, Perier MC, Singh-Manoux A, Gaye B, Thomas F, Prugger C, et al. Cross-sectional analysis of deprivation and ideal cardiovascular health in the Paris Prospective Study 3. Heart. 2016;102(23):1890–7.

52. Gupta B, Gupta R, Sharma KK, Gupta A, Mahanta TG, Deedwania PC. Low Prevalence of AHA-Defined Ideal Cardiovascular Health Factors Among Urban Men and Women in India. Glob Heart. 2015.

53. Ren J, Guo XL, Lu ZL, Zhang JY, Tang JL, Chen X, et al. Ideal cardiovascular health status and its association with socioeconomic factors in Chinese adults in Shandong, China. BMC Public Health. 2016;16:942.

54. Seron P, Irazola V, Rubinstein A, Calandrelli M, Ponzo J, Olivera H, et al. Ideal Cardiovascular Health in the southern cone of Latin America. Public health. 2018;156:132–9.

55. van Nieuwenhuizen B, Zafarmand MH, Beune E, Meeks K, Aikins AD, Addo J, et al. Ideal cardiovascular health among Ghanaian populations in three European countries and rural and urban Ghana: the RODAM study. Internal and emergency medicine. 2018;13(6):845–56.

56. Younus A, Aneni EC, Spatz ES, Osondu CU, Roberson L, Ogunmoroti O, et al. A Systematic Review of the Prevalence and Outcomes of Ideal Cardiovascular Health in US and Non-US Populations. Mayo Clinic proceedings. 2016;91(5):649–70.

57. Mujahid MS, Moore LV, Petito LC, Kershaw KN, Watson K, Diez Roux AV. Neighborhoods and racial/ethnic differences in ideal cardiovascular health (the Multi-Ethnic Study of Atherosclerosis). Health & place. 2017;44:61–9.

58. Shockey TM, Sussell AL, Odom EC. Cardiovascular Health Status by Occupational Group - 21 States, 2013. MMWR Morbidity and mortality weekly report. 2016;65(31):793–8.

59. Dragano N, Bobak M, Wege N, Peasey A, Verde PE, Kubinova R, et al. Neighbourhood socioeconomic status and cardiovascular risk factors: a multilevel analysis of nine cities in the Czech Republic and Germany. BMC Public Health. 2007;7(1):255.

60. Clark AM, DesMeules M, Luo W, Duncan AS, Wielgosz A. Socioeconomic status and cardiovascular disease: risks and implications for care. Nature Reviews Cardiology. 2009;6:712.

61. Ferguson TS, Younger N, Tulloch-Reid MK, Forrester TE, Cooper RS, Van den Broeck J, et al. Prevalence of the metabolic syndrome in Jamaican adults and its relationship to income and education levels. The West Indian medical journal. 2010;59(3):265–73.

62. Ferguson TS, Younger-Coleman Nom, Tulloch-Reid MK, Bennett NR, Rousseau AE, Knight-Madden JM, et al. Factors associated with elevated blood pressure or hypertension in Afro-Caribbean youth: a cross-sectional study. PeerJ. 2018;6:e4385.

63. Rosengren A, Hawken S, Ounpuu S, Sliwa K, Zubaid M, Almahmeed WA, et al. Association of psychosocial risk factors with risk of acute myocardial infarction in 11119 cases and 13648 controls from 52 countries (the INTERHEART study): case-control study. Lancet (London, England). 2004;364(9438):953–62.

64. Nguyen XT, Quaden RM, Wolfrum S, Song RJ, Yan JQ, Gagnon DR, et al. Prevalence of Ideal Cardiovascular Health Metrics in the Million Veteran Program. The American journal of cardiology. 2018;122(2):347–52.

